# Frequency and Predictors of Adolescent Worry for School Gun Violence In the United States: Findings from a Nationally Representative Study

**DOI:** 10.1101/2025.02.18.25322472

**Authors:** Danielle Kirkland, Tsung-chieh Fu, Debby Herbenick, Devon J. Hensel

## Abstract

**Purpose:** While school gun violence (SGV) incidents in the United States (U.S.) have risen sharply over the past decade, limited research has examined adolescent worries about SGV. We examined the frequency and correlates of SGV worry in a U.S. nationally representative sample of adolescents.

**Methods:** Data were from adolescent participants (14–17 years; N = 1017) in the 2022 National Survey of Sexual Health and Behavior, a nationally representative study of sexual health experiences of people in the U.S. SGV worry was a single 5-point item (not at all worried – extremely worried). We used both weighted descriptive statistics to examine SGV frequency and random intercept mixed effects ordinal regression to examine demographic and background impact on SGV worry.

**Results:** Nearly 75% of adolescents reported some degree of SGV worry; of these, one in five were very or extremely worried. SGV worry was significantly higher for adolescents in younger grades and among racial/ethnic minority youth, as well as for cisgender female and gender minority teens. Adolescents in higher income homes were less worried about SGV. Both teens living in metropolitan locations and teens who reported higher anxiety in the past two weeks noted higher SGV worry.

**Conclusions:** U.S. adolescents have a substantial level of worry about school gun violence. Structural interventions are needed to reduce SGV itself. Moreover, because there are detrimental long-term impacts of prolonged worry, targeted interventions are important for reaching those who are at greatest worry risk, including lower income, race/ethnic minority and gender minority teens.

**Implications and Conclusions:** Three out of every four adolescents in the United States worry about being a victim of school gun violence. Structural interventions to both reduce SGV itself and to reach adolescents at greatest risk for SGV worry are warranted.

## Background

Firearm death is currently the leading cause of death among children and adolescents in the United States (U.S.)^1^ School gun violence (SGV) is a prominent subset of these deaths. In recent years, the U.S. has had nearly 60 times as many school shootings as all other industrialized nations combined.^2^ Since the Columbine High School shooting in 1999, more than 383,000 students nationally have been exposed to SGV in K-12 institutions.^3^ In the past decade, there have been at least 1383 incidents of gunfire on school grounds nationally; 71.7% of these incidents have resulted in injury, and one-third have resulted in death.^4^ In 2024 alone, there were 267 youth wounded or killed from gunfire on K-12 property^5^ – this is a rate of nearly one young person *per day*.

Adolescents can have both direct (e.g., being a SGV victim and/or having a family member/friend be a SGV victim) and indirect (e.g., seeing SGV on social or print media, or participating in school drills) exposure to SGV.^6,7^ Direct exposure has a clear adverse impact on adolescent academic, physical and emotional well-being,^8-10^ and may create worry about future violence. Far more adolescents will be indirectly exposed to SGV, which may also create worry. SGV events are widely and graphically depicted on news and social media.^11^ Many teens additionally face daily reminders of SGV in the form of increased security measures (e.g., metal detectors, armed guards or school resource officers).^12^ Simulation or preparedness drills can expose students to graphic SGV elements like the use of fake blood or the shooting of guns with blanks or rubber bullets.^13^ Some youth will personally face different forms of SGV threats (e.g., social media posts, written plans and drawings, messages on the bathroom wall, rumors about attacks, and anonymous 911 calls).^14^ These ongoing experiences may therefore increase youth worry about SGV in their own school, even if such violence has never occurred in their own communities.^11^ A 2014 convenience sample of high school students in California echoes these findings, with one-third of participants citing feeling very or extremely concerned, worried or stressed about shootings or violence at their own school.^15^ A 2018 survey had similar findings, with more than half of US students reporting feeling fearful or somewhat or very worried about the possibility of a shooting happening at their school.^16^ These sentiments are exacerbated in racial/ethnic, gender and sexual minority teens17-20

In the current paper, we use U.S. nationally representative survey data to examine SGV worry among 14 – to 17-year-old adolescents. Our first objective was to assess the population level frequency and significance of self-reported SGV worry in this age group. The marked increase in SGV in the past three years justifies a closer look at the extent to which such an uptick may impact the prevalence and magnitude of teen worry about GGV. Our second objective was to examine the association of adolescent demographic and health characteristics with SGV worry. It is important to understand if some youth are at higher worry risk for worry as compared to others, as heightened states of negative emotion may create additional downstream physical and mental health challenges.

## Methods

### Study Design and Participants

Data were from the 2022 National Survey of Sexual Health and Behavior (NSSHB), a cross-sectional and U.S. nationally representative survey examining sexuality and well-being among individuals aged 14 and over years of age in the United States (U.S.). This study is the eighth iteration in a series of population based NSSHB surveys conducted since 2009. Data collection was facilitated by Ipsos Research (Menlo Park, California) using their KnowledgePanel® – a probability-based web panel of 60,000 adults designed to be representative of noninstitutionalized U.S. citizens over the age of 18 who can read and complete surveys in English or Spanish languages. The KnowledgePanel® was established using an address-based sampling (ABS) frame using the U.S. Postal Service’s Delivery Sequence File (e.g., the panel is established via probability methods and is not an “opt in” panel). ABS improves population coverage and enhances the ability to sample hard-to-reach individuals, such as young adults, those without landline telephones, and racial/ethnic minorities. KnowledgePanel® has been used for numerous U.S. nationally representative surveys on diverse topics, including physical activity (Supplemental Reference 1), dementia (Supplemental Reference 2), COVID-19 (Supplemental Reference 3) and sexual health(Supplemental References 4 and 5).

Sampled households are invited to join KnowledgePanel through a series of mailings, including an initial invitation letter, a reminder postcard, and follow up phone calls. Households that do not have internet access may be provided with a web-enabled device and free internet to facilitate participation. After initially accepting the invitation to join the panel, participants are asked to complete a short demographic survey; answers to this survey allow efficient panel sampling and weighting for future surveys. Upon completing the initial survey, adult participants become active panel members. All panel members are provided with privacy and confidentiality protections. Members typically receive no more than one survey invitation per week; on average they complete two to three surveys per month. Compensation is provided via a modest incentive program that includes raffles and sweepstakes with both cash rewards and other prizes for completing surveys. No additional compensations were given in this study.

We reached adolescents through their adult KnowledgePanel® member parents, beginning with a sampling frame of 3357 adult KnowledgePanel® members who were parents/guardians of 14–to 17-year-olds. During December 2022 and January 2023, Ipsos sent these adult members a message, either via email or through their online dashboard, that let them know a new survey was available. Those who clicked on the link were provided with detailed information about the study, which was described as being about people’s “sexual health and experiences” and being for all people “regardless of whether they have ever even kissed anyone or had sex.” Parents could electronically indicate their consent to participate and then proceed to complete the survey. Of the 3357 parents/guardians in the sampling frame, 43.2% (N = 1451) consented for their adolescent to be invited into the study. Of the 1451 adolescents contacted about the study, 1017 (70.1%) agreed to participate. Recruitment occurred fully online (e.g., at no point did our research team have any direct contact with participants) until the targeted number of complete surveys (*N* = 1000) was reached; at that point, Ipsos closed recruitment. The confidential online survey was offered in English and in Spanish and had a median completion time of 21 minutes.

Once the data were collected and processed, Ipsos provided post-stratification statistical weights to account for differential nonresponse (i.e., under-coverage or over-coverage of certain demographic groups). Weights were created using demographic distributions for based on population-specific demographic distributions from the 2021 Current Population Survey (CPS), and accounted for gender, Census region, Metropolitan status, household income, ethnicity, income, ethnicity, education, and language proficiency (English proficiency, Bilingual, Spanish proficiency, Non-Hispanic) by age based on the 2021 Current Population Survey (CPS). Once created, resulting weights are scaled to aggregate to the total sample size of all eligible respondents. Multiple weights were provided depending upon any oversampling or specific demographic groups targeted. For the current study, we used the adolescent specific weights. Ipsos then sent a de-identified data set to our research team. The Indiana University Institutional Review Board reviewed and approved protocols and measures associated with the NSSHB-8 (IRB Protocol #16792).

### Measures

#### SGV worry

Adolescents reported the extent to which they worried about “gun violence at their own, a family member’s or a friend’s school” (not at all, slightly, somewhat, very or extremely).

#### Predictor variables

Ipsos provided several parent-reported demographic variables, including *age* (years: 14, 15, 16 and 17), *race/ethnicity* (White, Non-Hispanic; Black, Non-Hispanic; Other, Non-Hispanic; Hispanic; and 2+ races, Non-Hispanic), *household size* (number of people), *annual household income* (recategorized to facilitate analyses: low income [<$10,0000 to $49,999], middle income [$50,000 - $99,999], upper-middle income [$100,000 - $149,000] and high income [≥$150,0000]) and *home location* (non-metropolitan vs. metropolitan). Participants also described their gender (man, woman, gender non-binary, transgender women or transfeminine, transgender man or transfeminine, or some other term). We trichotomized this information into *gender identity* (man, cisgender woman, gender minority [gender nonbinary, gender queer, transgender and those who used a different term). We additionally examined *sexual orientation identity* (heterosexual, gay or lesbian, bisexual, asexual and some other term) and *past two week anxiety* (Patient Health Questionnaire:^21^ low vs high anxiety).

##### Statistical Approach

To evaluate Objective #1, we used weighted frequencies to establish the population level frequency of self-reported SGV worry among the sample of adolescents. We then performed a Wilcoxon ranked sign test on the SGV worry variable to establish whether the reported worry levels were significantly different from not at worried – or “no worry” – at the population level. To evaluate Objective #2, we used random intercept mixed effects ordinal regression to evaluate the impact of demographic and health variables – as expressed by an odds ratio (OR) and corresponding 95% confidence interval (CI) – on SGV worry level. Intercepts were allowed to vary across states to adjust for clustering of adolescents within states with different gun restriction policies and volume of prior SGV. Preliminary unconditional random effects models intraclass correlation coefficient (ICC) analyses exceeded thresholds (ICC: 0.07; threshold: 0.05) for assumption of meaningful nesting of responses within states.^22^ All analyses were conducted in Stata, v. 18.0.

## Results

### Participants

Sample characteristics are shown in Table 1. The sample was an average age of 15.6 years and the majority (87.9%) were enrolled in high school, while 10% were in middle school. They predominantly described themselves as White, Non-Hispanic (51.4%), followed by Hispanic (24.8%), and Black, Non-Hispanic (13.1%). Most were men (50.7%) or women (45.6%); 3.4% identified as gender nonbinary, transgender, or other terms. A high percentage of adolescents identified as heterosexual (86.7%). The average parent-reported household size was 4.3 members, and the majority of participants lived in parent-reported metropolitan areas. The median parent-reported household income was middle-income ($50,000-$99,999). One in three adolescents (32.5%) were classified as having high anxiety risk.

**Table 1.** Demographic and background characteristics of a nationally representative sample of adolescents (N=1010) in the 2022 National Survey of Sexual and Health Behavior.

| Characteristics | Overall % (N)<br>Mean (SD) |
| --- | --- |
| SGV Worry Level |  |
| Not at all | 25.9 (261) |
| Slightly | 27.7 (278) |
| Somewhat | 27.9 (281) |
| Very | 12.3 (124) |
| Extremely | 6.1 (61) |
| Age | 15.6 (1.1) |
| 14 | 24.6 (248) |
| 15 | 24.2 (245) |
| 16 | 26.2 (265) |
| 17 | 25.0 (252) |
| Grade |  |
| Middle school (7<sup>th</sup> and 8<sup>th</sup> grade) | 10.0 (101) |
| High school (9<sup>th</sup> – 12<sup>th</sup> grade) | 87.3 (880) |
| Higher education (technical school or college) | 1.6 (16) |
| Not enrolled in school | 1.3 (13) |
| Race/ethnicity |  |
| White, Non-Hispanic | 51.4 (519) |
| Black, Non-Hispanic | 13.1 (132) |
| Other, Non-Hispanic | 6.4 (65) |
| Hispanic | 24.8 (250) |
| 2+ races, Non-Hispanic | 4.3 (44) |
| Gender identity |  |
| Cisgender man | 50.7 (512) |
| Cisgender woman | 45.6 (461) |
| Gender nonbinary, transgender or some other term | 3.4 (35) |
| Sexual identity |  |
| Gay or Lesbian (ref) | 5.1 (510) |
| Heterosexual | 86.7 (868) |
| Bisexual | 2.5 (25) |
| Pansexual | 2.8 (28) |
| Asexual | 0.9 (9) |
| Queer | 0.8 (8) |
| I use another term | 1.2 (12) |
| Household size | 4.3 (1.4) |
| Household income |  |
| Low income (<\$10,000 to \$49,999) (ref) | 30.8 (312) |
| Middle income (\$50,000 - \$99,999) | 34.1 (345) |
| Upper-middle income (\$100,000 - \$149,000) | 20.2 (204) |
| High income (≥\$150,000) | 14.8 (150) |
| Home location |  |
| Non-metro (ref) | 14.1 (143) |
| Metro | 85.9 (867) |
| Anxiety risk |  |
| Low risk | 67.4 (666) |
| High risk | 32.5 (988) |

### Objective 1 – Population-Level Prevalence and Significance of SGV Worry among Adolescents

Table 1 displays the weighted population prevalence of overall SGV worry levels. Wilcoxon rank sign test confirmed that these worry levels were significantly different from “not at all worried” (*t*=23.838; *p*<.001) – in other words, a significant proportion of adolescents in the U.S. were worried about gun violence at their own, a family member’s or a friend’s school. We found specifically that nearly *three out of four adolescents* (73.3%; 744/1011) reported this worry. Of these, about one in five reported high levels – either very (12.3%: 124/744) or extremely (6.1%: 61/744) – of being worried about SGV. About a third of adolescents reported being slightly (27.6% 278/744) or somewhat (281/744) worried about SGV.

### Objective 2 – Predictors of Adolescent SGV Worry

Weighted random intercept mixed effects ordinal regression results are shown in Table 2. Adolescents in older grades tended to report lower levels of SGV worry (OR=0.83); however, in supplemental models, comparing school stage (e.g. middle school, high school, some post-secondary education) did not yield significant differences. Being a racial/ethnic minority was associated with increased risk of SGV: adolescents who identified as Other/Non-Hispanic (OR=2.01) had two-fold higher SGV fear as compared to White/Non-Hispanic adolescents, and Hispanic adolescents reported three-fold higher SGV worry (OR=2.96) than White/Non-Hispanic adolescents. Both women (OR=1.38) and gender minority (OR=2.16) teens were significantly more worried about SGV than men teens. There was insufficient power to further investigate SGV worry between specific gender minority categories (e.g., gender nonbinary vs. transgender women or men).

**Table 2.**
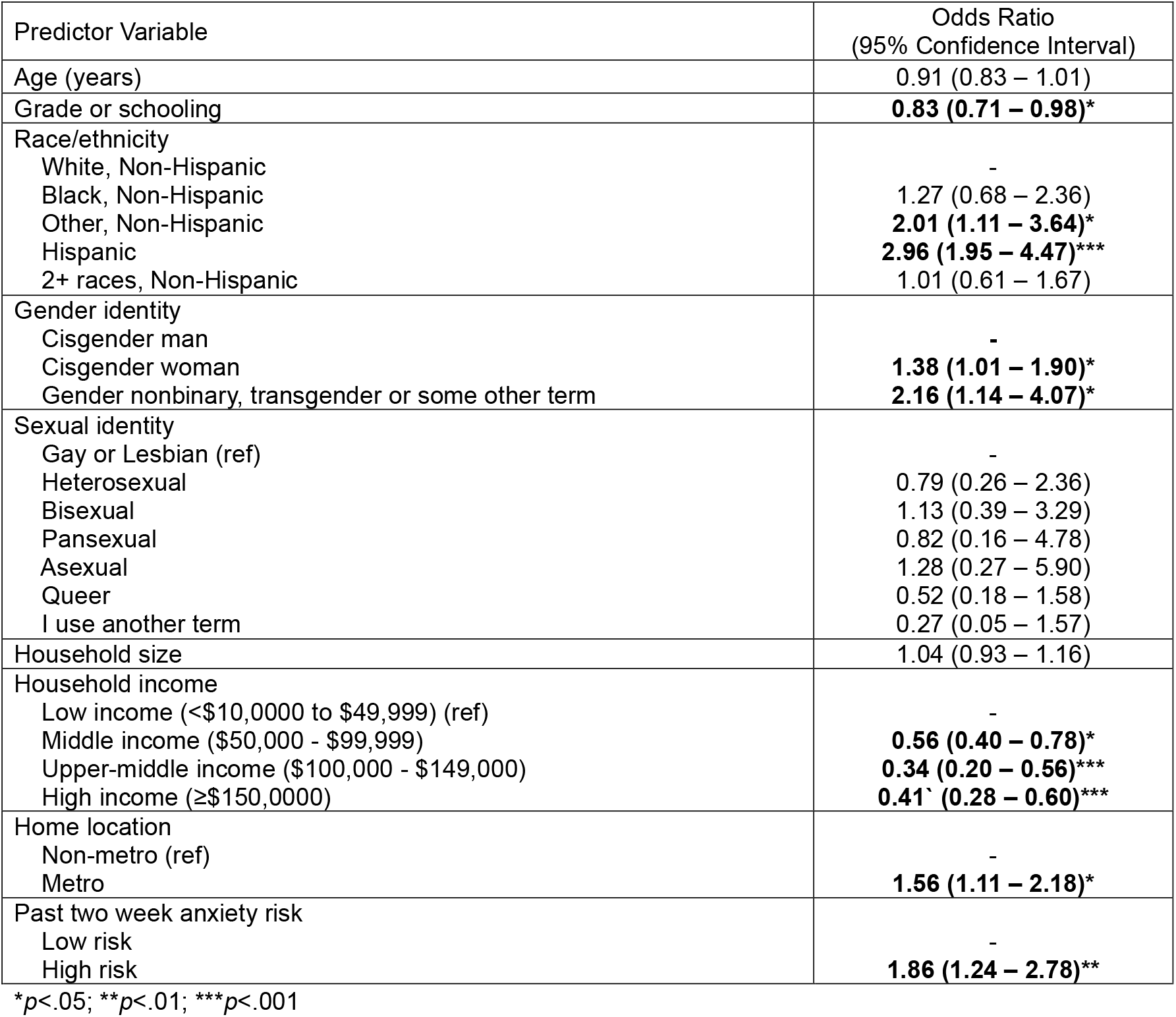
Mixed effect ordinal regression results – predictors of adolescent (N=1010) school gun violence worry in the 2022 National Survey of Sexual and Health Behavior.

SGV fear became significantly less as household income increased (OR=0.31 – 0.56), while adolescents living in metropolitan areas (OR=1.56) reported significantly higher odds of SGV than adolescents in non-metropolitan areas. Finally, higher risk of past two-week anxiety was associated with higher odds of SGV worry (OR=1.86). There was no significant association of age, school type, household size or sexual identity on adolescent SGV.

### Supplemental Models

We conducted supplementary models to investigate whether different predictor variable construction impacted our findings. Specifically, we created dichotomized versions of sexual orientation identity (heterosexual vs. gay/lesbian, bisexual, asexual, queer and something else) and race/ethnicity (White vs. Black, Other, 2+ races and Hispanic) to explore the possibility of interaction between minority statuses. We specified an alternative model included the main effect of these new variables, the main effect of gender identity, two-way interactions between new sexual identity and new race/ethnicity, between new sexual orientation identity and gender identity and between gender identity and new race/ethnicity and gender identity. This model also controlled for other original covariates. The main effect of race/ethnicity showed that adolescents of color (OR=2.39; *p*<.001) and women (OR=1.64; *p*=.006) reported higher SGV worry. Neither the two-way interaction between gender identity and sexual orientation identity (*p*=-0.351-0.569) nor between sexual orientation identity and race (*p*=0.18) were significant. A significant two-way interaction between gender identity and race/ethnicity suggested that gender diverse adolescents of color have higher SGV worry than cisgender male adolescents of color (OR=1.94; *p*=.014). Because the overlapping minority categories were small, we did not have enough statistical power to investigate these effects further. Such interactions should be a priority in future research.

## Discussion

The purpose of this study was to describe the percentage of U.S. adolescents who worry about school gun violence (SGV) and to understand which demographic or health characteristics may be associated with increased or decreased worry. Given the dire – and still worsening – epidemic of school shootings in this country,^1,3,7,23^ it should surprise no parent, clinician, educator or other youth-focused professional that adolescents in the U.S. also experience SGV worry at epidemic levels. Three out of every four adolescents in our sample reported they worried to some extent about gun violence at their school or a family member’s or a friend’s school. These rates exceed those in the few available studies in the extant research from 2014 (71.8%)^15^ and from 2018 (57%).^16^ Moreover, one of every five teens who worried reported that they did so at a high level. Longitudinal data using U.S. nationally representative samples are needed to further understand the extent to which the rate and/or severity of school shootings – or perhaps even perceptions of safety – impact how deeply adolescents are concerned about future violence.

Our findings also highlight important disparities in adolescent SGV worry. We found that Black Non-Hispanic and Other Non-Hispanic adolescents had two-to three-fold the odds of worry as compared to White adolescents. These data align with prior studies that demonstrate elevated fear, worry and/or concern about school gun violence among students of color.^16,24^ While our worry measure did not ask specifically about worry source, other work has suggested that youth of color are more likely to report being exposed to violence either at home or at school as compared to White peers,^25^ which may be linked to heightened anticipatory concern about being at future risk for gun violence.^26^ Our data additionally suggest that women and gender minority adolescents were significantly more worried about SGV than their men peers. While no prior studies have specifically examined SGV worry in gender minority youth, other research does suggest that women, transgender, gender expansive and questioning youth are in increased risk for different types of violence both at school and in relationships.^27-29^ Similarly to adolescents of color, these experiences likely heighten worry about gun violence. In alignment with previous studies, adolescents from lower income household reported higher SGV,^16^ perhaps because their exposure to violence near school is more likely than high income peers.^25^

Finally, we also found that adolescent at higher risk for anxiety also demonstrated higher SGV worry. It is now well understood that both direct and indirect exposure to school shooting is detrimental to children and adolescents’ mental health,^30,31^ and some longitudinal studies have shown that increased SGV worry is associated with increased anxiety and panic disorder.^15^ The cross-sectional nature of this study did not permit us to disentangle the causal order of SGV worry and anxiety risk, and while SGV worry may indeed predict higher anxiety, we think it is also important to consider the possibility that adolescents with existing mental health challenges may be pre-disposed to experience higher worry about school shootings. It will be important for future studies to employ designs capable of understanding how these affective trajectories impact one another over time.

While the risk factors for SGV worry likely extend beyond what we were able to examine in this study, we believe our data offer preliminary support for different interventions to reduce adolescent worry. One always important intervention is the implementation of sensible and consistent gun control laws to appropriately limit weapons access (e.g., criminal background checks, age-related restrictions, military-style assault weapons and high-capacity ammunition magazine bans, conceal and carry laws, reducing gun trafficking, required weapons registration, red-flag laws, etc.).^32^ The research is clear that stronger gun regulation is associated with decreased levels of gun-associated violence,^33^ and adolescents who live in states with tighter firearm laws are less likely to be threatened with weapons at school and are more likely to feel safe at school.^34^ However, creation of a federal culture of gun responsibility and safety will remain a challenge until weapons regulation is viewed as a pediatric public health matter rather than a political platform.

An additional intervention is the integration of mental health professionals in educational settings (e.g., school psychologists or social workers). These individuals provide a crucial point of intervention by providing mental and emotional care for students who may not otherwise have access to care [Supplemental References 5 and 6]. Adults in these role may be able to proactively identify and support healthy solutions for students whose own situation (e.g., anxiety or depression, substance use, family disruption, peer bullying) may either increase their SGV worry level or increase their tendencies towards any type of violence. Mental health personnel can also serve to foster a safe and connected school climate. Research suggests that SGV is significantly lower in schools in which students feel a part of a community that supports them.^35^ Because It is likely that mental health professionals’ roles will continue to evolve as schools integrate them into a larger prevention plan, it is vital that SGV curriculum be included as part of their formal educational training and as part of professional development.^36^

Finally, educational environments and services themselves may serve as an intervention for worry reduction. As described earlier in this paper, many institutions increasingly employ daily SGV prevention (e.g., metal detectors, armed guards or officers, “zero tolerance” policies that suspend or expel students for violence threats or bullying) and preparedness (e.g., lockdown drills or ““run, hide, or fight” skills) strategies.^13^ Despite best intentions, many of these approaches have not actually decreased school shootings and/or other forms of violence,^30^ and may not significantly impact students’ perceptions of safety.^37^ These strategies often increase adolescent exposure to law enforcement in and around schools,^38^ which can be particularly traumatic for adolescents of color who already have increased anxiety and fear about such individual compared to their White peers.^39^ It is imperative for all schools and universities to carefully weigh the efficacy of implementing prevention and preparedness policies with their potential adverse impact on student well-being.

### Limitations

As with any study, some limitations should be considered in the interpretation of our results. First, as KnowledgePanel® members are adults, we had to obtain parental consent to recruit their adolescent children, which may have affected which teens had the option to participate. Second, the cross-sectional structure of the survey means that we were unable to establish causal order between variables of interest. Thus, while we assumed that adolescents with increased anxiety could be predisposed to increased SGV worry, it is also possible that worry itself exacerbates adverse mental health outcomes. In addition, survey length and concerns about participant burden prevented us from being able to ask more in depth questions about SGV experience, such as whether or not a participant was had ever personally been a victim of SGV and/or the extent to which they experienced any school based SGV prevention and preparedness strategies. Third, while we did find evidence of SGV worry clustering within states, we were unable to include a comprehensive measure that adequately operationalized those experiences – for example, gun control policies, volume of past and current school shootings. Fourth, as a U.S. nationally representative survey, our study is also limited by small numbers of sexual and gender minority participants; subsequent research is needed that oversamples individuals with these identities.

## Conclusion

School gun violence – and the worries and fears that adolescents have about it – is an important pediatric health issue. While most young people will not personally be involved in a violence event, their day-to-day lives are rife with reminders about the potentiality of victimhood. Using U.S. nationally representative data, we found that most adolescents have a substantial worry about SGV happening in their lives. In addition to implementing structural interventions to reduce SGV itself, targeted efforts to help adolescents at greatest risk for SGV worry are important to avoid the long-term mental health impacts of prolonged worry.

## Data Availability

All data produced in the present study are available upon reasonable request to the authors.

## Abbreviations

SGV: school gun violence

